# Mindfulness-Based Cognitive Therapy for Treatment-Resistant Depression: A protocol for a systematic review and meta-analysis

**DOI:** 10.1101/2024.06.16.24308994

**Authors:** Michele F Rodrigues, Larissa Junkes, Jose C Appolinario, Antonio E Nardi

## Abstract

**Background:** Major Depressive Disorder is a global health issue that affects more than 300 million people worldwide, which is about 4.4% of the global population. Treatment usually consists of a combination of antidepressant medication and therapy. However, approximately 30% of individuals with Major Depressive Disorder experience a Treatment-Resistant Depression (a failure to respond to at least two antidepressants used in a regular dosage and time interval). Our research aims to investigate how effective Mindfulness-Based Cognitive Therapy is for individuals with Treatment-Resistant Depression.

**Materials and Methods:** We will conduct a thorough search for publications in MEDLINE, Embase, PsycINFO, Web of Science databases, and ClinicalTrials.gov. Additionally, we will manually review the reference lists of the included studies to find any other potentially relevant studies. There will be no restrictions on language or publication date. The quality of the included studies will be assessed independently using the Jadad scale (Oxford quality scoring system). To assess the certainty of the evidence, we will use the Grading of Recommendations Assessment, Development, and Evaluation (GRADE) methodology. This study aims to determine the effectiveness and tolerability of Mindfulness-Based Cognitive Therapy as an intervention for Treatment-Resistant Depression.

**Ethics and dissemination:** Ethical approval is not necessary because individual patient data is not being collected. Our objective is to publish the systematic review in an open-access medical journal once the review process is completed.

**Prospero registration ID:** CRD42023411978. Registered on April 07, 2023.

## Introduction

Major Depressive Disorder (MDD) is a significant global health concern that affects over 300 million individuals worldwide, which is approximately 4.4% of the global population, according to estimates from the World Health Organization [1]. This condition is characterized by persistent feelings of a depressed mood or loss of interest that last for at least two weeks. Symptoms may also include anhedonia, changes in appetite and sleep patterns, feelings of worthlessness, low self-esteem, and difficulty concentrating [2]. Treatment typically involves a combination of antidepressant medication and therapy. However, a significant number of individuals, estimated to be between 29% and 46%, do not see improvement even after receiving the correct doses and duration of treatment. These individuals are categorized as having Treatment-Resistant Depression (TRD). [3]. Recent research has shown that 69.2% of individuals with Treatment-Resistant Depression do not achieve a satisfactory response even after one year of treatment. The remission rate is low, with 60% of individuals still receiving the same treatment as when the study began [4].

From a psychosocial perspective, individuals with Major Depressive Disorder and Treatment-Resistant Depression frequently experience negative emotions and exhibit dysfunctional thought patterns, such as persistently fixating on pessimistic thoughts. This ongoing cycle of anxiety and self-criticism can exacerbate the condition. When traditional medication fails to produce results, individuals may begin to lose hope of recovery, leading to pessimistic attitudes about their symptoms improving. Consequently, they may shy away from activities that could challenge these negative beliefs and aid in alleviating symptoms of anhedonia. Numerous studies have emphasized the beneficial impact of mindfulness in treating individuals with TRD by targeting these key areas of dysfunction [5,6].

Mindfulness is the awareness that arises by intentionally paying attention in the present moment, without judgment [7]. Originally, the practice of mindfulness meditation in Buddhism was intended to alleviate suffering and promote compassion [8]. Mindfulness-Based Cognitive Therapy (MBCT) combines mindfulness meditation from the Mindfulness-Based Stress Reduction (MBSR) protocol [7] with elements of Cognitive Behavioral Therapy (CBT) for depression [9]. MBCT can be administered in individual or group sessions, either in person or online, and typically lasts for eight weeks. The program aims to improve awareness of the present moment by practicing mindfulness meditation. This helps individuals identify and distance themselves from negative automatic thoughts and unhelpful attitudes [10]. It promotes new methods for recognizing and breaking free from harmful patterns of repeated negative thinking, while also fostering a nonjudgmental understanding of depressive thoughts and emotions. This helps to decrease symptoms of depression in TRD [11,12]. Significant reductions in the severity of depression have been shown between the groups that received the MBCT intervention and the control groups [11,13]. Moreover, the Cladder-Micus study investigated the effectiveness of combining MBCT with Treatment as Usual (TAU) for individuals who have chronic depression and are resistant to treatment. The study’s analysis revealed that although the intent-to-treat approach did not result in a decrease in depressive symptoms, individuals who completed MBCT+TAU experienced a notable reduction in depressive symptoms compared to those who only received TAU.

As there is strong evidence supporting the effectiveness of mindfulness interventions in various mental health settings, particularly for depression, this systematic review aims to address an important gap in the current literature. While there have been positive results for MBCT in reducing depressive symptoms in the general population, its effectiveness in individuals with

Treatment-Resistant Depression has not been well-studied. Therefore, this study aims to thoroughly review and analyze the existing evidence on the impact of MBCT on individuals diagnosed with TRD.

### Research question

How effective is Mindfulness-Based Cognitive Therapy when used in conjunction with standard pharmacological treatment for individuals with Treatment-Resistant Depression?

#### Objectives

To assess how individuals with Treatment-Resistant Depression respond when Mindfulness-Based Cognitive Therapy is added to their regular medication treatment, taking into account the following factors:

##### Effectiveness

###### Reduction in Depression Severity

A significant change in depression severity over the course of the treatment using standardized scales such as the Hamilton Depression Rating Scale (HDRS) [14], Inventory of Depressive Symptomatology-Self-Report (IDS-RS) [15] or the Beck Depression Inventory (BDI) [16].

###### Remission Rates

Evaluate the proportion of individuals achieving remission, defined as a significant reduction or absence of depressive symptoms or specified criteria validated in standardized depression scales.

###### Response Rates

Determine the proportion of individuals showing a clinically significant improvement over the course of the treatment in depressive symptoms using standardized scales for depression.

##### Psychological and Cognitive Outcomes

###### Mindfulness Skills

Five Facet Mindfulness Questionnaire (FFMQ) [17], assesses five facets of mindfulness: observing, describing, acting with awareness, non-judging of inner experience, and non-reactivity to inner experience. Southampton Mindfulness Questionnaire (SMQ) [18], measures the ability to respond mindfully to distressing thoughts and images.

###### Rumination

Ruminative Response Scale (RRS) [19], assesses the extent to which individuals focus on their depressive symptoms and the causes and consequences of those symptoms.

###### Self-compassion (SCS) [20], measures self-compassion across three dimensions

self-kindness versus self-judgment, common humanity versus isolation, and mindfulness versus over-identification.

##### Long-term Outcomes

###### Sustained Remission

Assess the sustainability of remission and symptom improvement over an extended follow-up period using standardized scales for depression.

###### Relapse Rates

Determine the rates of relapse and time to relapse after the intervention using standardized scales for depression.

## Materials and methods

### Study design

The protocol for this review was registered in the International Prospective Register of Systematic Reviews (PROSPERO) on April 7, 2023 (registration number PROSPERO 2023 CRD42023411978). This systematic review will adhere to the Preferred Reporting Items for Systematic Reviews and Meta-Analyses Protocol (PRISMA-P) guidelines [21].

### Eligibility criteria

We will search the MEDLINE, Embase, PsycINFO, Web of Science databases, and ClinicalTrials.gov. Additionally, we will manually review the reference lists of the included studies to find any relevant additional studies. There will be no restrictions based on language or publication date. To refine and structure our research question, we used the PICO framework. This framework defines our population as individuals with Treatment-Resistant Depression (TRD), our intervention as Mindfulness-Based Cognitive Therapy (MBCT), and our outcomes as primary outcomes related to depressive symptoms and secondary outcomes including quality of life and mindfulness skills. When applicable, the comparison group will include standard care or no intervention. This comparison will help evaluate the effectiveness of MBCT in a comparative context.

### Search strategy

The following primary search strategy will be used for MEDLINE: ((“Depressive Disorder, Major”;[Mesh] OR (Major Depressive Disorder) OR (Paraphrenia, Involutional) OR Involutional Paraphrenia*) OR (Paraphrenias, Involutional) OR (Psychosis, Involutional) OR (Involutional Psychos*) OR (Psychoses, Involutional) OR (Depression, Involutional) OR (Involutional Depression) OR (Melancholia, Involutional) OR (Involutional Melancholia)) OR (“Depressive Disorder, Treatment-Resistant”[Mesh] OR (Depressive Disorder*, Treatment Resistant) OR (Disorder*, Treatment-Resistant Depressive) OR (Treatment-Resistant Depressive Disorder*) OR (Refractory Depression*) OR (Depression*, Refractory) OR (Therapy-Resistant Depression) OR (Depression*, Therapy-Resistant) OR (Therapy Resistant Depression) OR (Therapy-Resistant Depressions) OR (Treatment Resistant Depression*) OR (Depression*, Treatment Resistant) OR (Resistant Depression*, Treatment))) AND (“Mindfulness”[Mesh]).

### Selection criteria

In order to be included in this review, articles must meet the following criteria: (1) Participants must be adults (18 years or older) diagnosed with TRD; (2) The MBCT intervention must be compared with either active treatments (such as medications or other forms of psychotherapy) or inactive treatments (such as placebos or waiting lists); (3) The primary outcome, which is the improvement in depression symptoms, must be measured using established and validated instruments; (4) Participants must have a confirmed diagnosis of both Depression and Treatment-Resistant Depression; (5) The review will consider study designs such as Randomized Controlled Trials (RCTs), follow-up studies derived from RCTs, quasi-experimental studies, and systematic reviews; (6) Grey literature will be included through manual searches and reviews of reference lists to identify additional sources; (7) If MBCT is combined with other interventions, these will be evaluated separately; (8) Studies involving individuals under 18 years of age will be excluded; (9) Participants with bipolar disorder or substance abuse issues will not be considered; and (10) Studies with incomplete protocols will be excluded.

### Studies selection

Two reviewers (MFR and LJ) will each independently screen search results based on titles and abstracts using predefined criteria to evaluate eligibility and eliminate any duplicate entries according to the specified inclusion criteria for the review. The selection results will be compared, and any disagreements regarding eligibility will be resolved through consensus or, if necessary, by consulting a third party (AEN). Study selections and reasons for exclusion will be carefully recorded to create a comprehensive PRISMA flowchart [21]. If multiple publications from the same study are encountered, data will be selected from the most comprehensive outcomes. In cases of duplicates, the most recent publication will be utilized.

### Data extraction

Two independent reviewers (MFR and LJ) will use a form developed specifically for this review to extract data from the final selected studies. If necessary, the authors will be contacted to obtain additional relevant or missing information. The fields to be extracted from eligible studies include the author’s name, publication date, study design, socio-demographic data, diagnostic criteria for Treatment-Resistant Depression, type of intervention, and scales used for assessing depression severity. Data extraction will focus on primary outcomes related to depressive symptoms, secondary outcomes such as quality of life and mindfulness skills, and any reported adverse effects. The extracted data will also include information on the duration and frequency of MBCT sessions, participant adherence rates, and any follow-up assessments conducted by the studies. This comprehensive approach ensures a thorough evaluation of MBCT’s effectiveness and safety for individuals with TRD.

### Quality assessment

For the review study, only the randomized clinical trials will undergo evaluation of quality and risk of bias. To control for bias, the guidelines of the Jadad scoring system [22] will be used, with two authors (MFR and LJ) independently applying the inclusion criteria. One of these authors will be responsible for extracting the data and assessing its quality, while the other reviewer will verify these results. Any differences between their assessments will be resolved through consensus. The Jadad scoring system assesses several key aspects: randomization, random allocation concealment, masking of treatment allocation, blinding, and withdrawal. Outcome selection bias will be assessed by comparing reported outcomes to published results. Intention-to-treat analysis will be used, with the population consisting of randomized participants who attend at least one MBCT session and are evaluated post-baseline.

### Data analysis

In our systematic review, we aim to combine the data using both quantitative and qualitative analyses, depending on the nature and compatibility of the data extracted from the included studies. Our approach is as follows:

### Quantitative Synthesis

If the data allows, we will conduct a meta-analysis to evaluate the effectiveness of Mindfulness-Based Cognitive Therapy in reducing symptoms of Treatment-Resistant Depression. This will involve combining data from studies that have utilized similar interventions, comparators, and outcome measures. We will use a random-effects model to address between-study variations, which will be assessed using the chi-square test or by calculating I^2^. Heterogeneity will be considered low or absent if I^2^ is less than 50%, while I^2^ equal to or greater than 50% will be considered significant heterogeneity. If there is no statistical heterogeneity (I^2^ less than 50% and P value greater than 0.1), we will use the fixed-effects model for meta-analysis. In cases of statistical heterogeneity (I^2^ equal to or greater than 50% and P value less than or equal to 0.1), the random-effects model will be utilized for meta-analysis.

Continuous outcomes will be analyzed utilizing standardized mean differences with Hedges’ g for effect size calculation. Dichotomous outcomes will be analyzed using risk ratios, although we expect the outcomes will primarily be continuous. We will investigate potential sources of heterogeneity through subgroup analyses and meta-regression, with a focus on variables such as study quality, participant demographics, and intervention characteristics. To assess the robustness of our findings, sensitivity analysis will be performed by excluding studies with a high risk of bias. Publication bias will be evaluated utilizing funnel plots, the trim-and-fill method, and classic fail-safe N calculations, as appropriate.

### Qualitative Synthesis

For studies that provide qualitative data or are not suitable for meta-analysis, we will conduct a narrative synthesis. This will involve extracting and summarizing details on the methodologies, themes, and concepts that are relevant to our research questions from each study. Any discrepancies in data extraction and interpretation will be resolved through discussions among reviewers, and a structured summary will be produced for each article. The data will be tabulated to aid in comparing across studies. We will integrate the qualitative and quantitative findings into a comprehensive narrative synthesis, following the guidance [23]. The decision on the specific methods of synthesis will be made iteratively, based on the available data, and the rationale for the chosen methods will be documented and reported.

### Software and Tools

For the quantitative synthesis, we will utilize Comprehensive Meta-Analysis Version 3 [24] and R software version 3.1.1 (Comprehensive R Archive Network) [25] for statistical analyses, which includes calculating effect sizes, evaluating heterogeneity, and assessing publication bias. The findings will be visually presented through forest plots. As for the qualitative synthesis, the data will be structured and analyzed using NVivo software [26] to aid in recognizing and summarizing themes across studies.

This two-pronged approach will enable us to gain a comprehensive understanding of how effective MBCT is in treating depression that is resistant to conventional treatment. We will consider both the quantifiable results and the subtle insights gathered from qualitative data. In order to evaluate the reliability of the findings, we will utilize the Grading of Recommendations Assessment, Development, and Evaluation (GRADE) methodology. This methodology assesses five key criteria: risk of bias, imprecision, inconsistency, indirect evidence, and publication bias.

### Ethics and Dissemination

As this is a systematic review and meta-analysis, ethical approval is not necessary. The findings from this review will be published in a peer-reviewed journal and will also be presented at appropriate conferences.

## Discussion

Given the significant impact and enduring high rates of depression over the years, it is crucial to conduct a comprehensive and up-to-date review of available treatment methods. There have been meaningful progress in pharmacological treatments that have introduced new ways to alleviate the debilitating effects of the disease. Enhanced interventions are necessary to address this problem. Combining psychotherapy with pharmacological treatment has the potential to bring positive results. However, there is limited evidence on which psychotherapeutic techniques are most effective for individuals with Treatment-Resistant Depression (TRD). The expected outcomes may show that the effectiveness of Mindfulness-Based Cognitive Therapy combined with antidepressant medication in TRD is better than pharmacotherapy alone. This planned review and meta-analysis will carefully examine the available evidence for TRD and the role of MBCT in this subgroup. By gathering and summarizing information, this study aims to improve our understanding of existing gaps and identify the most promising approaches to enhance patient response. Additionally, the results of this review will guide future research efforts and provide valuable information for healthcare practitioners.

## Data Availability

No datasets were generated or analysed during the current study. All relevant data from this study will be made available upon study completion.

## Supporting information

**S1. Checklist. PRISMA-P (Preferred Reporting Items for Systematic review and Meta-Analysis Protocols) 2015 checklist: Recommended items to address in a systematic review protocol\***

Corresponding author: Michele Ferreira Rodrigues, Institute of Psychiatry (IPUB) of Federal University of Rio de Janeiro (UFRJ), Venceslau Braz Avenue, 71, Botafogo, Rio de Janeiro – RJ, 22290-140, Brazil. Email:

Larissa Junkes

Jose Carlos Appolinario Antonio E. Nardi

## Notes

### Competing Interest Statement

The authors have declared no competing interest.

### Funding Statement

The author(s) received no specific funding for this work.

### Author Declarations

This study does not require ethical approval because it involves a systematic review and meta-analysis of previously published studies. No individual patient data is being collected or analyzed. Therefore, no Institutional Review Board (IRB) or oversight body approval is necessary for this research. Details of the IRB/Oversight Body: Not applicable.

